# Genomic Epidemiology of Healthcare-Associated Respiratory Virus Infections

**DOI:** 10.1101/2025.04.20.25325828

**Authors:** Vatsala Rangachar Srinivasa, Marissa P. Griffith, Alexander J. Sundermann, Emma Mills, Nathan J. Raabe, Kady D. Waggle, Kathleen A. Shutt, Tung Phan, Anna F. Wang-Erickson, Graham M. Snyder, Daria Van Tyne, Lora Lee Pless, Lee H. Harrison

**Affiliations:** Microbial Genomic Epidemiology Laboratory, Center for Genomic Epidemiology, University of Pittsburgh, Pittsburgh, PA, USA; Division of Infectious Diseases, University of Pittsburgh School of Medicine, Pittsburgh, PA, USA; Department of Epidemiology, School of Public Health, University of Pittsburgh, Pittsburgh, PA, USA; Department of Infection Control and Hospital Epidemiology, UPMC, Pittsburgh, PA; Department of Pediatrics, Division of Infectious Diseases, University of Pittsburgh School of Medicine, Pittsburgh, PA, USA; Institute of Infection, Inflammation, and Immunity in Children (i4Kids), Pittsburgh, PA; Department of Pathology, University of Pittsburgh School of Medicine, Pittsburgh, PA, USA

**Keywords:** whole genome sequencing, respiratory viruses, genomic epidemiology, healthcare-associated infections, outbreak

## Abstract

**Background:** Respiratory virus transmission in healthcare settings is not well understood. To investigate the transmission dynamics of common healthcare-associated respiratory virus infections, we performed retrospective whole genome sequencing (WGS) surveillance at one pediatric and two adult teaching hospitals in Pittsburgh, PA.

**Methods:** From January 2, 2018, to January 4, 2020, nasal swab specimens positive for rhinovirus, influenza, human metapneumovirus (HMPV), or respiratory syncytial virus (RSV) from patients hospitalized for ≥3 days were sequenced on Illumina platform. High-quality genomes were assessed for genetic relatedness using ≤3 single nucleotide polymorphisms (SNPs) cut-off, except for rhinovirus (10 SNPs). Patient health records were reviewed for genetically related clusters to identify epidemiological connections.

**Results:** We collected 436 viral specimens from 359 patients: rhinovirus (n=291), influenza (n=50), HMPV (n=47), and RSV (n=48). Of these, 55% (197/359 patients) were from pediatric hospital and 45% from adult hospitals. Patients ranged in age from 14 days to 93 years, 61% were male, and 74% were white. WGS was performed on 61.2% (178/291) rhinovirus, 78% (39/50) influenza, 92% (44/48) RSV, and all HMPV specimens. Among high-quality genomes, we identified 14 genetically related clusters involving 36 patients, ranging in size from 2-5 patients. We identified common epidemiological links for 53% (19/36) of clustered patients; 63% (12/19) patients had same-unit stay, 26% (5/19) had overlapping hospital stays, and 11% (2/19) shared common provider. On average, genetically related clusters spanned 16 days (range:0−55 days).

**Conclusion:** WGS offered insights into respiratory virus transmission dynamics. These advancements could potentially improve infection prevention and control strategies, leading to enhanced patient safety and healthcare outcomes.

**Summary:** We performed retrospective whole genome sequencing surveillance of common healthcare-associated respiratory virus infections across three hospitals. Our investigation elucidated complex respiratory virus transmission dynamics, which could potentially improve infection prevention and control strategies, leading to enhanced patient safety and healthcare outcomes.

## Introduction

Acute respiratory infections result in approximately 4.25 million deaths globally each year [1]. In 2017, influenza was associated with approximately 54 million cases and 145,000 deaths [2]. Respiratory syncytial virus (RSV) alone is estimated to cause 14,000 deaths annually among U.S. adults over 65, with a global burden of around 64 million infections and 160,000 deaths annually [3]. Respiratory virus infections are especially severe in older adults, immunocompromised individuals, and children under five [1, 2, 4, 5]. In the United States, respiratory viruses such as influenza, human metapneumovirus (HMPV), and RSV are predominantly seasonal, often circulating between October through April, whereas rhinovirus is present year-round [4].

Healthcare-associated respiratory virus infections lead to longer hospital stays, higher morbidity, and increased healthcare resource utilization, such as ICU admissions and mechanical ventilation, leading to higher costs [1, 6–9]. The hospital population often comprises young children, older adults, immunocompromised patients, and those with underlying health conditions, who are at heightened risk of severe respiratory virus infections [10–12]. Despite this, transmission of common respiratory viruses in hospital settings is understudied, mostly focusing on single-species transmission [10–19]. This indicates a significant gap in knowledge regarding infection control and patient safety.

Traditional epidemiological methods are commonly used to track and manage the spread of respiratory viruses. These methods typically involve monitoring for significant increases in infection incidence above baseline levels, which then trigger follow-up investigations [20–22]. This approach limits the ability to determine outbreaks as soon as a single transmission event, which is necessary for implementing timely interventions and for effectively controlling the spread of the virus early in an outbreak [23]. Traditional methods also fail to distinguish between a single outbreak or multiple outbreaks of different viral variants.

Recently, the addition of whole genome sequencing (WGS) has been used to augment traditional outbreak investigation, known as reactive WGS [12, 14–16]. Traditional outbreak investigations complemented with reactive WGS could delay outbreak identification and miss outbreaks entirely. We performed retrospective WGS surveillance of healthcare-associated respiratory viral infections, including rhinovirus, influenza, RSV, and HMPV, to further understand their transmission dynamics and as a pilot study for possible future implementation of real-time WGS surveillance.

## Methods

### Study setting

This study was performed in three teaching hospitals in Pittsburgh, PA, two primarily serving adults and one pediatric hospital, each having a total bed capacity exceeding 700 beds. Ethics approval was obtained from the University of Pittsburgh Institutional Review Board (STUDY21040126).

### Specimen collection

From January 2, 2018, to January 4, 2020, viral transport media of anterior nasal swab specimens positive for respiratory viruses, including rhinovirus, influenza, RSV, and HMPV from the three study hospitals were collected, deidentified, and stored at –80°C until further processing. During this period, respiratory virus testing was done at the discretion of the clinical provider. Specimens were included for inpatients who had been hospitalized for ≥3 days and/or had a recent inpatient or outpatient encounter within 30 days before the positive test date.

### Specimen processing

Total nucleic acid was extracted using the MagMAX Viral and Pathogen Nucleic Acid Isolation Kit (ThermoFisher Scientific) per manufacturer’s instructions. Quantitative real-time polymerase chain reaction (qPCR) was performed for all specimens using technical duplicates to identify the cycle threshold (Ct) value. We used TaqMan microbe detection assay (ThermoFisher Scientific) for RSV and HMPV, CDC’s influenza/SARS-CoV-2 multiplex assay for influenza [24], and a SYBR green assay for rhinovirus [25]. The qPCR data were analyzed by manually adjusting the threshold to approximately three-quarters of the log-exponential phase. Specimens with Ct values ≤30, <32, and ≤39 for rhinovirus, influenza, and RSV, respectively, were prepared for WGS per manufacturer’s directions (Twist Bioscience).

Sequencing libraries were generated using one of two methods. For all rhinovirus, influenza, and RSV Ct ≥24, a hybridization method was used (respiratory virus research panel, Twist Bioscience) [26]. We modified how we combined individual rhinovirus libraries into one pool, prior to the 16-hour hybridization step; these libraries were pooled based on viral copy number instead of equimolar pooling (**Supp Methods**). Each rhinovirus library pool contained eight specimens. The libraries for RSV and influenza specimens were pooled at an equimolar concentration based on Ct values (approximately +/− 2 SD); each pool contained between four and eight specimens. Additionally, during protocol optimization, eight rhinovirus and two influenza specimens with Ct values higher than our cut-off values were sequenced.

For all HMPV specimens, regardless of the Ct value, and for RSV <24 Ct, a tiled PCR amplicon method followed by Illumina DNA Prep was used (**Supp Methods**) [27, 28]. For HMPV, apart from the primers described by Tulloch et al.,2021 an additional PCR 4 forward primer (5’-GGTCATAAACTCAAAGAAGGTG-3’) was designed to enhance amplification. All libraries were sequenced on the Illumina platform.

### Bioinformatics analyses

Viral species were determined using Kraken2 (v2.1.2) [29], followed by the removal of reads mapping to the human genome. The resulting reads were assembled using the RNA viral SPAdes (v3.15.5) *de novo* approach with default parameters [30], except for influenza, which was assembled using IRMA (v1.0.3) [31]. For viral species other than influenza, contigs were then aligned to a reference genome chosen based on nucleotide similarity from a blastn search of publicly available genomes in NCBI; scaffolds were generated using nucmer (v4.0.0) [32]. CheckV (v.1.0.1) was used to estimate the completeness of the *de novo* assembly [33]. Viral subtypes were determined using NCBI blastn. Genomes passed WGS QC if ≥90% of the genome had ≥10× coverage depth and had at least 90% estimated completeness from CheckV.

Assembled genomes were aligned using Multiple Alignment using Fast Fourier Transform (MAFFT, v7.515) [34]. We trimmed the 5’ and the 3’ areas of the multiple genome alignment to ensure there were no missing bases in those regions and to eliminate sequencing bias due to a drop in sequencing depth in these regions of the genome. The resulting alignment file was used to determine single nucleotide polymorphisms (SNPs) and to obtain a maximum-likelihood phylogenetic tree using a generalized time-reversible (GTR) approach with FastTree (v0.11.2), with default parameters [35]. The phylogenetic tree was visualized with iToL [36]. To identify genetically related transmission clusters, a pairwise SNP analysis using SNP-sites (v2.5.1) [37], followed by distance-based hierarchical clustering, was performed. Based on our prior experience investigating viral outbreaks in hospital settings, a 3-SNP cut-off with average linkage clustering was chosen to define genetically related clusters of influenza, RSV, and HMPV. To account for the higher mutation and diversity in rhinovirus [38], a 10-SNP cut-off was used for the whole genome analysis.

### Partial genome analysis

To assess hospital transmission, prior studies have often analyzed partial genomes of influenza and rhinovirus, instead of the full genome [11, 12, 15]. For influenza, these studies focused on the variable hemagglutinin (HA) segment; for rhinovirus, the internal ribosomal entry site (IRES) spanning approximately the first 1000 bases of the viral genome is often used. For influenza and rhinovirus, we investigated the utility of these regions for transmission if the genomes had >90% coverage at ≥10× depth using a 3– or 2-SNP cut-off for clustering, respectively.

### Epidemiological analyses

We reviewed patient electronic health records to identify common epidemiological links among patients in genetically related clusters. We defined common epidemiological link as: 1) same hospital unit stay, 2) shared provider, or 3) overlapping hospital stay. A shared unit implied shared healthcare providers. The shared provider category indicated that patients were not on the same unit but were attended by the same healthcare provider.

## Results

During the study period, we collected 436 rhinovirus (n = 291), influenza (n = 50), HMPV (n=47), and RSV (n=48) specimens from 359 patients (**Table 1**). Of the 359 unique patient respiratory virus specimens, 152 (42.34%) were from the pediatric hospital (PH), 124 (34.54%) from adult hospital 1 (AH1), and 83 (23.12%) from adult hospital 2 (AH2). The majority of patients were male (n=209, 58.2%) and white (n=267, 74.4%). There were 48 patients who had more than one respiratory virus specimen collected (range: 2-12; median, 2), and five patients had more than one respiratory virus identified.

**Table 1.** Demographic characteristics of patients with collected respiratory virus specimens.

|  |  | **Pediatric hospital,**<br>**N (%)**<br>**N = 152 (42.3)** | **Adult hospital 1,**<br>**N (%)**<br>**N = 124 (34.5)** | **Adult hospital 2,**<br>**N (%)**<br>**N = 83 (23.1)** |
| --- | --- | --- | --- | --- |
| **Sex** | **Female** | 54 (35.5) | 60 (48.4) | 36 (43.4) |
|  | **Male** | 98 (64.5) | 64 (51.6) | 47 (56.6) |
| **Race** | **White** | 111 (73) | 99 (80) | 57 (68.7) |
|  | **Black** | 20 (13) | 20 (16) | 21 (25.3) |
|  | **Other** | 6 (4) | — | 2 (2.4) |
|  | **Unspecified** | 15 (10) | 5 (4) | 3 (3.6) |
| **Mean Age** |  | 4.2 | 55.2 | 64.3 |
|  |  | Range 14 days-39 years<sup>#</sup> | Range 21-89 years | Range 25-93 years |
| **Respiratory**<br>**Season\*** | **2017-2018** | 38 (25) | 23 (18.6) | 27 (32.5) |
|  | **2018-2019** | 70 (46) | 84 (67.7) | 42 (50.6) |
|  | **2019-2020** | 44 (29) | 17 (13.7) | 14 (16.9) |
| **Organism** | **Rhinovirus/Enterovirus** | 116 (76.3) | 56 (45.2) | 51 (61.5) |
|  | **Influenza A Virus** | 3 (2) | 28 (22.6) | 12 (14.5) |
|  | **Influenza B Virus** | 3 (2) | — | 4 (4.8) |
|  | **RSV A** | 9 (5.9) | — | 2 (2.4) |
|  | **RSV B** | 11 (7.2) | 18 (14.5) | 5 (6) |
|  | **HMPV** | 10 (6.6) | 22 (17.7) | 9 (10.8) |

\* The duration of specimen collection varied each season: 2017 – 2018: 6 months; 2018 – 2019: 12 months; 2019 – 2020: 6 months; <sup>#</sup> Adults with complications since childhood continue to receive care at the children's hospital. Dash indicates the absence of specimens/patients in the specified category.
RSV, respiratory syncytial virus; HMPV, human metapneumovirus

Of all 436 viral specimens that underwent qPCR, 178 (61.2%) rhinovirus, 39 (78%) influenza, and 43 (90%) RSV specimens passed the qPCR QC criteria, respectively (**Supp Table 1**). The median Ct value for rhinovirus specimens was 27.5 (range:16.2-41), influenza was 26.7 (range:17-45), RSV was 28.8 (range:18.72-45), and HMPV was 30.7 (range:20.3-39.5) (**Supp Figure 1**). We obtained high-quality draft genomes for 114/178 (64%) rhinovirus/enterovirus, 34/39 (87%) influenza, and 41/43 (95%) RSV specimens. Additionally, 4/8 (50%) rhinovirus and 1/2 (50%) influenza specimens with high Ct values passed WGS QC. Nearly all (37/38, 97.4%) HMPV specimens with Ct values ≤37 passed WGS QC, suggesting that a higher Ct cut-off could potentially be used to perform WGS on HMPV specimens. qPCR was unable to distinguish between rhinovirus and enterovirus due to high genetic similarity, leading to the inclusion of two enterovirus specimens into our sequencing workflow; however, we identified these genomes using the WGS data and subsequently removed them from further analyses. Two RSV genomes had subtypes that did not match those identified by qPCR and were removed. Additional details on successfully sequenced genomes are provided in **Supp Table 5**.

**Figure 1.**
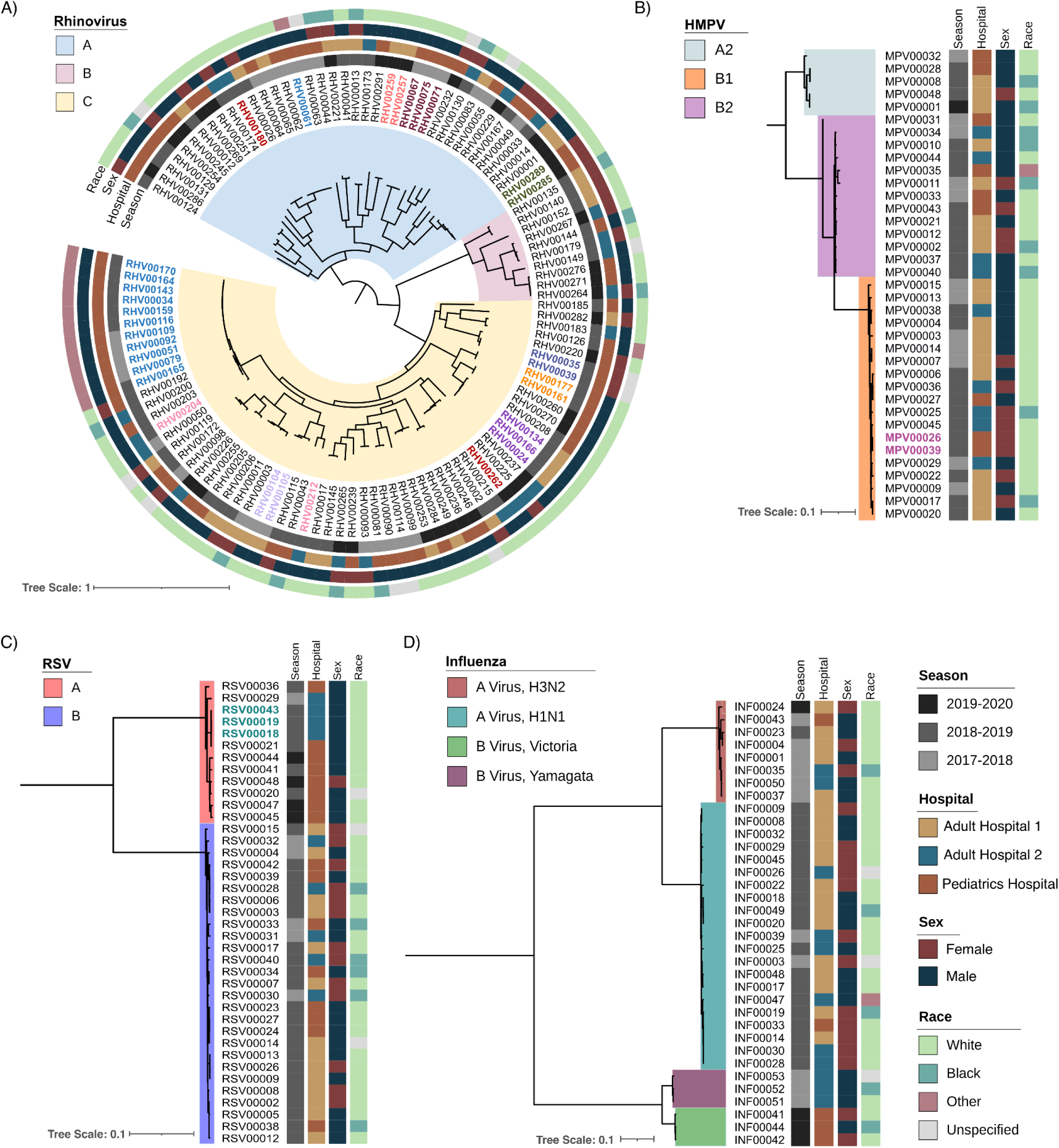
Phylogenetic tree of rhinovirus, human metapneumovirus (HMPV), respiratory syncytial virus (RSV), and influenza, by respiratory season, hospital, sex, and race. Tree scale represents the nucleotide substitutions per site. Colored branches represent viral subtypes,

We determined the subtypes of rhinovirus, HMPV, and influenza A and B from viral genomes that passed WGS QC (**Figure 1**). The majority of rhinovirus genomes belonged to subtype A (n=64), followed by subtypes C (n=41) and B (n=10), with one rhinovirus genome being most similar to a publicly available genome with no subtype reported. For influenza A, there were 21 H1N1 and eight H3N2 genomes and, for influenza B, there were three genomes each of the Yamagata and Victoria lineages. For HMPV, the majority of the genomes belonged to subtype B1 (n=20), followed by B2 (n=12), and A2 (n=5).

We next assessed whether there was sufficient genetic diversity to determine if WGS could reliably identify putative transmission events. The same-subtype pairwise SNPs for RSV ranged between 0–186 SNPs (median=91), HMPV ranged between 0–250 SNPs (median=114), and influenza within the same subtype ranged between 0–277 SNPs (median=136; **Figure 2**). Rhinovirus had the widest range of pairwise SNPs among genomes belonging to the same subtype, ranging from 0–2305 SNPs (median=1934; **Figure 3a**). This suggests that the viral populations sampled were genetically diverse and, therefore, appropriate for the analysis of closely related genomes.

**Figure 2.**
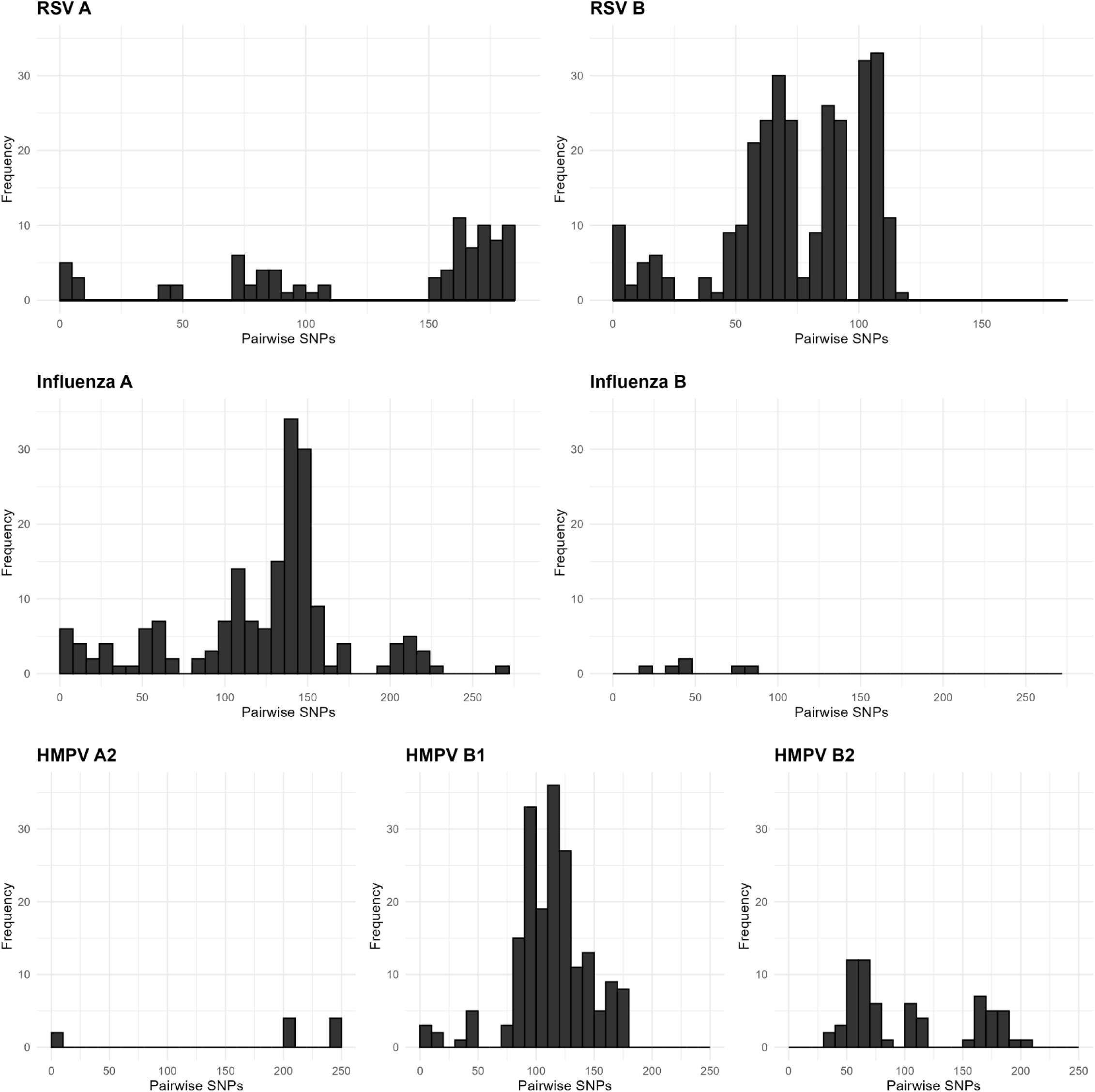
Pairwise single nucleotide polymorphism (SNP) distributions for RSV (respiratory syncytial virus), influenza, and HMPV (human metapneumovirus) genomes. Pairwise SNPs were assessed for all genomes in a given viral subtype, and histograms show the distribution of pairwise SNP distances.

**Figure 3.**
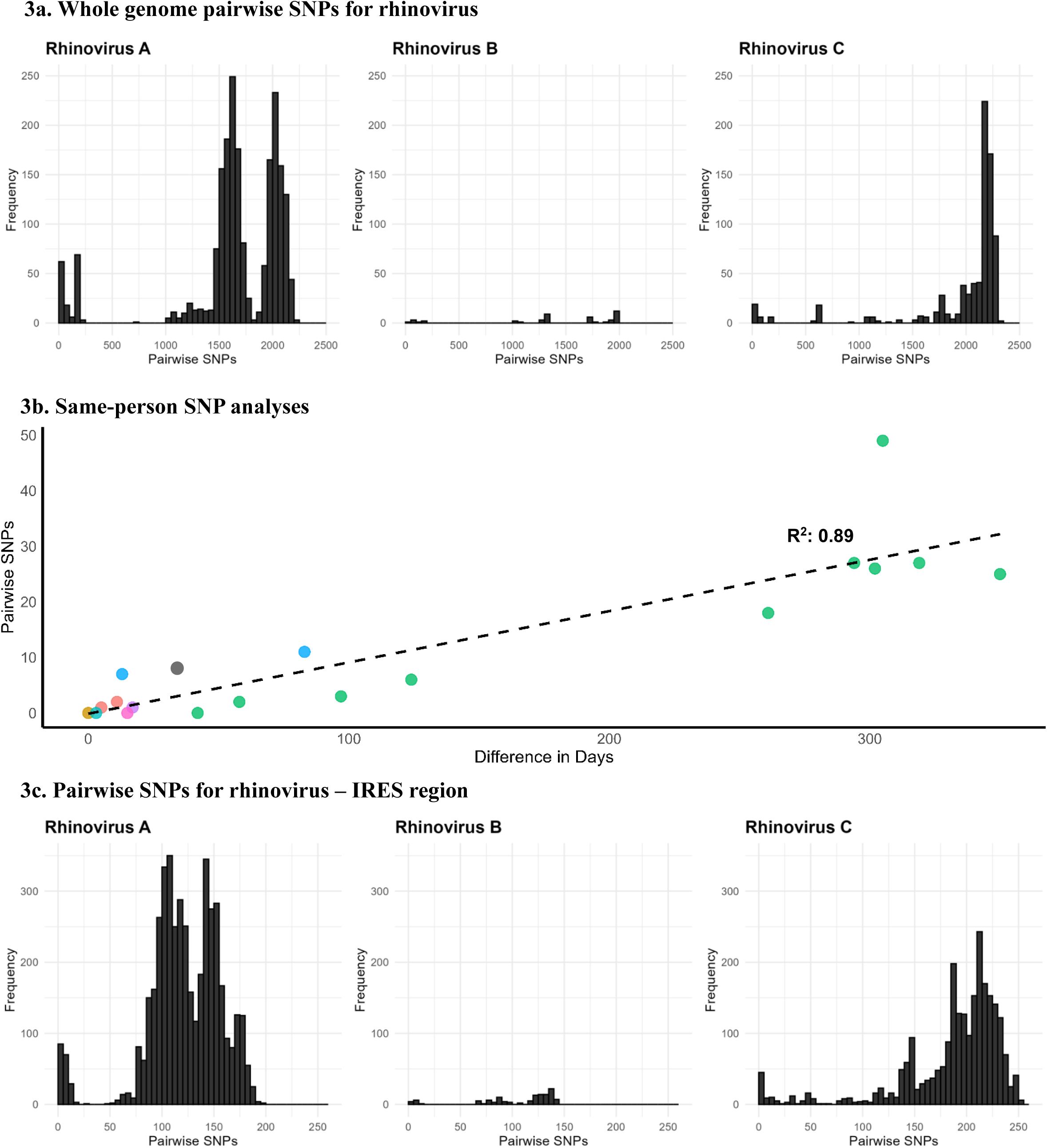
**3a**. Whole genome pairwise SNP distributions for different rhinovirus subtypes; **3b.** Pairwise SNPs vs. days between specimens for rhinovirus specimens collected from the same patients and belonging to the same subtype (an outlier with >1500 SNPs was removed from the plot). Circles of the same color represent individual

We identified 14 genetically related clusters containing 36 patients, ranging from 2–5 patients per cluster (**Table 2**); no patients were involved in more than one cluster. Overall, 36/359 (10%) patients were included in a transmission cluster. Rhinovirus genomes formed 6 clusters, followed by RSV and HMPV with 3 clusters each, and 2 clusters of influenza (**Figure 4**). We identified common epidemiological links in 19/36 (53%) patients. Of these, 12/19 (63%) patients shared the same unit prior to or at the time they tested positive, 5/19 (26%) had an overlapping hospital stay, and 2/19 (11%) shared a common healthcare provider. As a more detailed example, RSV cluster 7 (**Figure 5)** showed two distinct possibilities of transmission. First, patients 1 and 3 were exposed to RSV in unit 5, with the possibility of patient 3 having a longer incubation period, transmitting to patient 2 on a separate unit (unit 7). Second, patients 1 and 2 had out-of-unit exposure, with patient 2 transmitting the virus to patient 3 during the same unit stay (unit 7). Additionally, a fourth patient at a different hospital also had the same virus, suggesting complex transmission dynamics. In RSV cluster 9, all three patients had an overlapping same unit stay (unit 10), indicating likely exposure from that location. Patients from multiple hospitals were identified in 9/14 (64%) clusters. On average, the duration of genetically related clusters was 16 days, ranging from 0–55 days.

**Figure 4.**
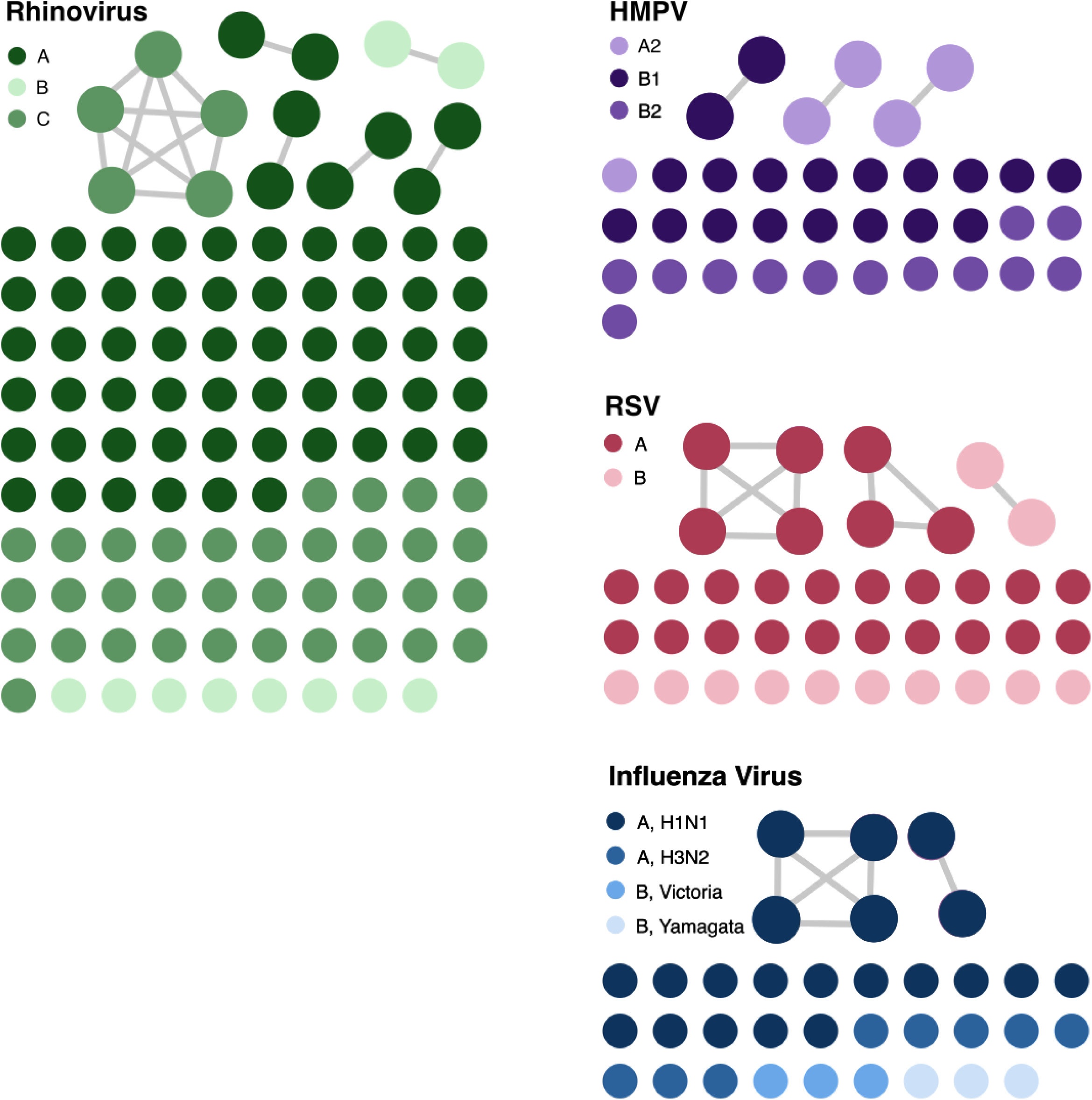
Cluster networks of respiratory virus genomes analyzed for putative transmission. The different color gradients within each virus represent different subtypes of the virus. The connected circles show patient specimens that were genetically related as defined by cutoffs described in the Methods section. The network plot was visualized with Gephi.

**Figure 5.**
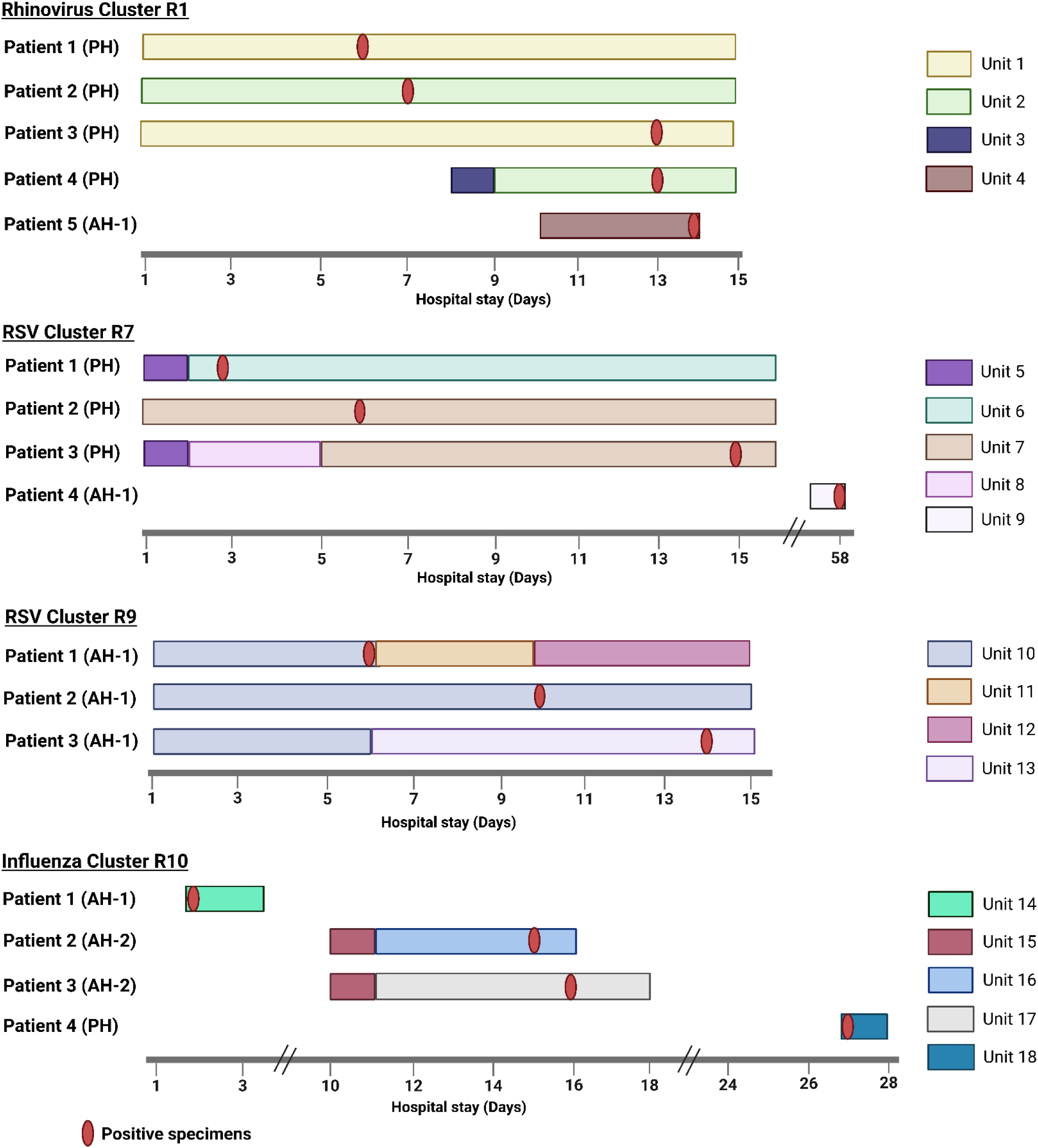
Presumed transmission patterns of genetically related respiratory virus clusters with >2 patients. Day 1 on x-axis is 5 days before the first positive test date within a cluster unless the specimen was collected on the first day of admission. Created in BioRender. Rangachar Srinivasa, V. (2025) https://BioRender.com/mdnle2e

**Table 2.** Summary of genetically related clusters.

| Cluster | No. Patients | Hospital | Epidemiologic link | Virus | Time between first and last specimen (days) |
| --- | --- | --- | --- | --- | --- |
| 1 | 5 | PH (n=4)<br>AH-1 (n=1) | PH: Overlapping stay | Rhinovirus C | 8 |
| 2 | 2 | PH | Shared unit | Rhinovirus A | 12 |
| 3 | 2 | PH (n=1)<br>AH-2 (n=1) | Unknown | Rhinovirus A | 12 |
| 4 | 2 | PH (n=1)<br>AH-2 (n=1) | Unknown | Rhinovirus A | 0 |
| 5 | 2 | PH (n=1)<br>AH-1 (n=1) | Unknown | Rhinovirus A | 2 |
| 6 | 2 | PH (n=1)<br>AH-2 (n=1) | Unknown | Rhinovirus B | 5 |
| 7 | 4 | PH (n=3)<br>AH-1 (n=1) | PH: Shared unit<br>AH-1: Unknown | RSV B | 55 |
| 8 | 2 | PH (n=1)<br>AH-2 (n=1) | Unknown | RSV A | 33 |
| 9 | 3 | AH-1 | Shared unit | RSV B | 8 |
| 10 | 4 | PH (n=1)<br>AH-1 (n=1)<br>AH-2 (n=2) | AH-2: Shared unit<br>PH/AH-1: Unknown | Influenza A | 25 |
| 11 | 2 | AH-1 | Shared unit | Influenza A | 4 |
| 12 | 2 | PH (n=1)<br>AH-1 (n=1) | Unknown | HMPV A2 | 0 |
| 13 | 2 | AH-1 | Unknown | HMPV A2 | 32 |
| 14 | 2 | PH | Common provider | HMPV B1 | 9 |

PH, Pediatric hospital; AH-1, Adult hospital 1; AH-2, Adult hospital 2; RSV, respiratory syncytial virus; HMPV, human metapneumovirus

A total of 29 rhinovirus genomes from nine patients were obtained through repeated sampling of the same patient (2–11 genomes per patient). Pairwise SNP analysis of these same-person, same-subtype genomes were closely related to one another (median: 6, range: 0-49 SNPs), with SNPs increasing as the time between specimen collection advanced (**Figure 3b**). In one patient, two rhinovirus A specimens that were collected 10 days apart, differed by 1,541 SNPs, suggesting possible co-infection with different strains of rhinovirus A. Additionally, there were two HMPV specimens from the same patient, collected 45 days apart and differed by 1 SNP, and three RSV specimens from one patient collected over 31 days that were genetically identical. Together, these data demonstrate the ability to identify both co-infection and prolonged infection using WGS data.

We explored using partial genomes for influenza and rhinovirus to assess transmission, as previously published [11, 12, 15]. For influenza, with a 3 SNP cut-off for the HA segment, the whole genome SNPs ranged from 0–34. For rhinovirus, pairwise SNPs in the IRES region ranged from 0–254 (**Figure 3c**). A 0 SNP cutoff in the IRES region corresponded to 0–52 SNPs in the whole genome, while a 1 SNP cutoff corresponded to 7–63 SNPs. These findings suggest that using only the HA segment for influenza and IRES for rhinovirus to define transmission was inadequate.

## Discussion

In this study, we demonstrated that WGS surveillance could provide valuable insights into the transmission patterns of rhinovirus, influenza, RSV, and HMPV in three Pittsburgh hospitals. This study adds to the existing literature by using systematic WGS surveillance for healthcare-associated infections of four different respiratory viruses. We successfully sequenced 74% of the genomes that passed the qPCR Ct criteria, identifying 14 genetically related clusters of respiratory viruses with 36 patients. The overall proportion of patients in a transmission cluster was 10%. A plausible transmission route was identified for 53% of these patients.

Conversely, we were unable to identify common epidemiological links for 47% of the patients with genetically related genomes. It is possible that healthcare workers (HCWs) or visitors were a source of transmission; thus, we were unable to identify the link because we did not have these specimens to sequence. Moreover, our inability to identify a common link could be due to the fact that there simply was no epidemiologic connection. At the time of this study, there was no requirement for regular asymptomatic screening of HCWs or universal masking. Therefore, we could not determine if patients were in contact with respiratory virus positive HCWs. It is also possible that some of these patients might have been in direct contact with each other in common hospital spaces, such as the cafeteria or the play area in the pediatric hospital. Additionally, 64% of clusters included patients from multiple hospitals, suggesting the possibility of community transmission, such as transmission at common gatherings, places of employment, daycare centers, or similar settings [39]. Furthermore, this could also be due to under-diagnosis of respiratory infections and non-sequenced positive specimens. However, investigating these additional scenarios was beyond the scope of this study. Future real-time WGS surveillance studies could focus on interviewing patients when a cluster is identified to better understand complex transmission dynamics.

The average duration of clusters was 16 days, ranging between 0–55 days, suggesting the possibility of longer contagious periods and possible asymptomatic transmission. Future investigations should focus on assessing the duration of contact and droplet precautions followed in hospitals for patients with respiratory virus infections. These long-spanning clusters would be undetected via traditional epidemiological investigations, highlighting the importance of WGS surveillance in identifying cryptic transmission.

Interestingly, one immunocompromised patient sampled in this study tested positive for rhinovirus for approximately one year (**Figure 3b**, green data points). One of the rhinovirus genomes sequenced from this patient, collected 16 days after the initial positive swab, was genetically related to viruses sampled from other patients. This suggests prolonged infection with intra-host viral evolution and transmission to other patients, consistent with other reports of immunocompromised patients with prolonged respiratory virus shedding serving as a source of transmission [40–42].

Relying on the genetic relatedness of only the HA segment for influenza and IRES region for rhinovirus has been a common practice in identifying influenza and rhinovirus transmission [11, 12, 15]. Our analyses showed that the partial genome SNPs for influenza and rhinovirus underestimated the genome-wide SNPs and resulted in overcalling transmission and was insufficient to confirm or refute transmission. Furthermore, a 10 SNP cut-off was used for whole genome analyses of rhinovirus transmission. However, no epidemiological links were found beyond a 3 SNP difference, suggesting that this cut-off is sufficient for identifying rhinovirus transmission clusters.

Our study has several limitations. First, we were not able to sequence all the collected specimens because some did not pass sequencing QC criteria. Second, we might have underestimated the number of transmission clusters due to our inclusion criteria and/or the arbitrary SNP cut-offs used. Third, we might have missed identifying common epidemiological links, such as a shared provider between two patients, due to limitations associated with the review of patient electronic health records. Also, not every HCW-patient encounter was reported in the electronic health records. Fourth, since this was a retrospective study, we could not evaluate the implications of our findings on infection prevention and control practices. This makes it difficult to understand whether real-time WGS surveillance of respiratory viruses would be useful in reducing transmission.

In conclusion, the integration of WGS surveillance and review of electronic health records elucidated transmission patterns across three hospitals. While this approach shows potential for detecting healthcare-associated transmissions of respiratory viruses, its broader implications warrant additional investigation. These advancements have the potential to improve infection prevention and control strategies, leading to enhanced patient safety and healthcare outcomes. Future studies should focus on performing real-time WGS surveillance of respiratory viruses to determine the potential utility of this approach for routine infection prevention and control practice.

## Data Availability

All data produced in the present work are either contained in the manuscript or NCBI/GISAID IDs are provided in a supplementary table.

## Acknowledgments

We are grateful to the UPMC Clinical Microbiology/Virology lab for helping with respiratory virus specimen collection. We also appreciate Dr. Jane Marsh’s support in optimizing the viral sequencing protocol.

## Data Sharing Statement

The NCBI and GISAID accession numbers for the respiratory viral genomes used in this study can be found in **Supp Table 5**.

## Author Contributions

VRS: Conceptualization, Methodology, Analysis, Data Curation, Writing – Original Draft, Writing – Review & Editing, Visualization

MPG: Data Curation, Analysis, Writing – Review & Editing

AJS: Methodology, Analysis, Data Curation, Writing – Review & Editing

EGM: Data Curation & Visualization, Writing – Review & Editing

NJR: Data Curation & Visualization, Writing – Review & Editing

KDW: Data Curation, Writing – Review & Editing

KAS: Data Curation, Writing – Review & Editing

TP: Methodology, Writing – Review & Editing

A.F.W.-E: Methodology, Writing – Review & Editing

GMS: Conceptualization, Writing – Review & Editing

DVT: Conceptualization, Methodology, Supervision & Writing – Review & Editing

LLP: Conceptualization, Methodology, Analysis, Project administration, Supervision & Writing – Review & Editing

LHH: Conceptualization, Methodology, Analysis, Resources, Data Curation, Writing – Original Draft, Writing – Review & Editing, Supervision, Project administration, Funding acquisition

## Supplementary material

**Supplementary Table 1.** Overview of whole genome sequencing metrics.

| Metrics/virus | Rhinovirus | Influenza A/B | RSV A/B | HMPV |
| --- | --- | --- | --- | --- |
| **Collected** | 291 specimens | 50 specimens | 48 specimens | 47 specimens |
| **Passed qPCR QC** | 178/291 specimens<br>(61.2%) | 39/50 specimens<br>(78%) | 43/48 specimens<br>(90%) | — |
| **Passed WGS QC** | 114/178 genomes<br>(64%) | 34/39 genomes<br>(87%) | 41/43 genomes<br>(95%) | 37/47 genomes<br>(79%) |
| **Failed species QC** | 2 genomes | — | 2 genomes | — |
| **High Ct genomes,**<br>**passed WGS QC\*** | 4/8 genomes<br>(50%) | 1/2 genomes<br>(50%) | — | — |
| **Total genomes**<br>**included (whole**<br>**genome analyses)** | 116 genomes | 35 genomes | 39 genomes | 37 genomes |
| **Genetically related** | 15 patients<br>(6 clusters) | 6 patients<br>(2 clusters) | 9 patients<br>(3 clusters) | 6 patients<br>(3 clusters) |
| **Epidemiologically**<br>**linked** | 7/15 patients<br>(47%) | 4/6 patients<br>(67%) | 6/9 patients<br>(67%) | 2/6 patients<br>(33%) |

\* While optimizing the protocols for whole genome sequencing, we sequenced few high Cycle threshold specimens of rhinovirus and influenza. Dash indicates the absence of respiratory viral specimens/genomes in the specified category.
RSV, respiratory syncytial virus; HMPV, human metapneumovirus

**Supplementary Figure 1.**
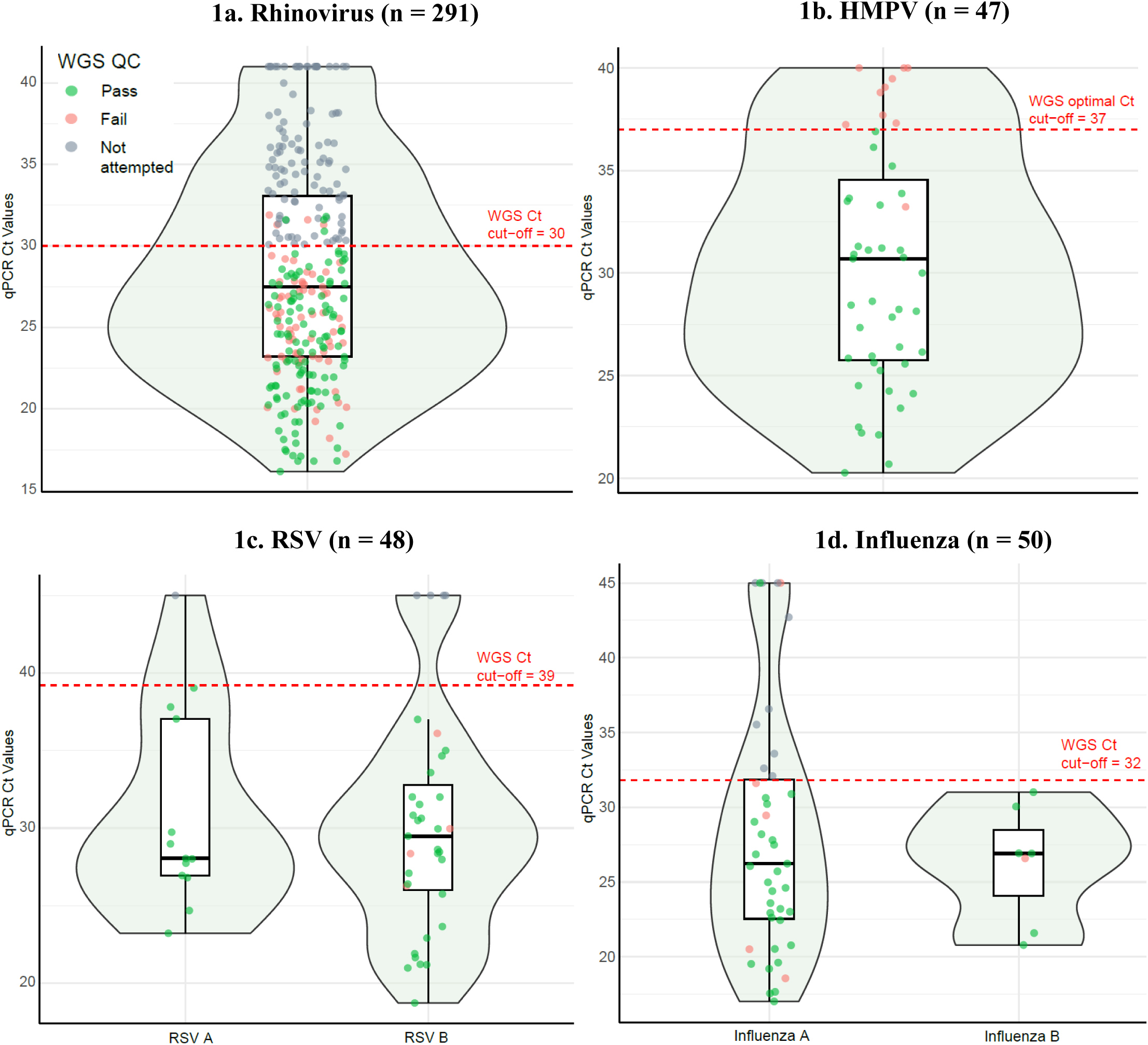
qPCR cycle threshold values (Ct) value for each of the respiratory viruses sequenced. The red dashed line indicates the qPCR Ct cut-off to perform whole genome sequencing (WGS); all HMPV specimens underwent WGS, and sequencing results indicated that a qPCR Ct cutoff of 37 is optimal for successful WGS. The dots within the box plot indicate the WGS QC status. RSV, respiratory syncytial virus; HMPV, human metapneumovirus

## Supplementary Methods

### Copy number assay for Rhinovirus – for equimolar pooling of libraries

The manufacturer’s protocol for pooling specimens was to combine eight specimens into a single sequencing pool; however, we observed a notable difference in the number of reads per specimen in a given pool, where a specimen with a minimal to modest difference in Ct value to another specimen would result in an imbalance in the number of reads following sequencing. To overcome this limitation, we modified the protocol to pool specimens by copy number, instead of equimolar pooling. To quantify the viral copy number of the clinical rhinovirus specimens, we constructed a standard curve by serially diluting RNA obtained from a reference sample (quantitative genomic RNA from human rhinovirus 77 strain 130-63; ATCC; serial dilutions spanned 20×10^3^ to 20×10^-1^ copies/μl). A two-step qRT-PCR protocol was performed on these serially diluted standards. First, RNA was converted to cDNA using the New England Biolabs protoscript reverse transcriptase for the first and second strand cDNA synthesis. This was followed by SYBR green PCR amplification of cDNA with primers specific to rhinovirus as described in Ng. et al., 2016 (**Supp Table 2**)[1]. The resulting Ct values were used to construct a standard curve, which was utilized to determine the viral load of each clinical specimen (**Supp Table 3**).

**Supp Table 2.**
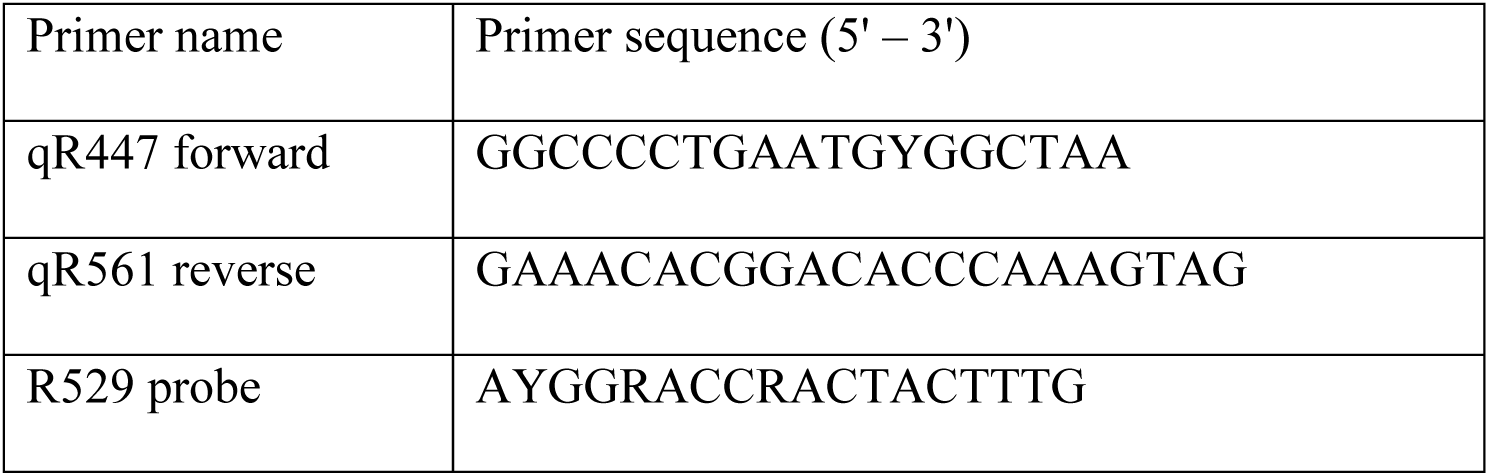
Primers used for rhinovirus qRT-PCR as described in Ng. et al., 2016.

**Supp Table 3.** Copy number calculation for rhinovirus.

| Quantity<br>(copies/μl) | Cycle threshold (Ct) |
| --- | --- |
| 20000 ($20 \times 10^3$) | 21.0 |
| 2000 ($20 \times 10^2$) | 24.3 |
| 200 ($20 \times 10^1$) | 27.6 |
| 20 ($20 \times 10^0$) | 31.8 |
| 2 ($20 \times 10^{-1}$) | 35.3 |

### Human metapneumovirus (HMPV) and respiratory syncytial virus (RSV) sequencing protocol modifications

For HMPV cDNA synthesis and genome amplification, we used a tiled single-plex PCR amplicon approach as described by Tulloch et al., 2021 that we modified to amplify the viral genomes prior to preparing the specimens for sequencing [2]. The cDNA synthesis was performed using SuperScript IV VILO Master Mix per manufacturer’s protocol (Invitrogen, Carlsbad, CA, USA). The cDNA was divided into four tiled, long-range PCR reactions, each separately amplifying one section of the HMPV genome. PCR amplification was performed using Platinum™ SuperFi™ PCR Master Mix per manufacturer’s protocol (Invitrogen). Apart from the primers mentioned in Tulloch et al. (**Supp Table 4**), an additional PCR 4 forward primer (5’-GGTCATAAACTCAAAGAAGGTG-3’) was designed to enhance amplification for some specimens that failed using the originally published primer.

Similar to HMPV, a tiled amplicon approach was used to amplify the RSV genome. A one-step reaction including cDNA synthesis and PCR amplification was used according to Dong et al., 2023 [3]. Six amplicons were combined into two primer pools that amplified the viral genome (**Supp Table 4**). The extracted total nucleic acid was divided into two reactions, one for each primer pool. We used SuperScript IV One-Step RT-PCR (Invitrogen) for both cDNA synthesis

For both HMPV and RSV, the resulting PCR product was visualized for quantification and to confirm amplification at the expected basepair size using the D5000 reagents on Agilent 4200 TapeStation system per manufacturer’s protocol. Specimens that successfully amplified were pooled into one tube to a final volume of 40 μl; we combined each pool based on concentration to ensure even sequencing coverage across the entire genome. The pooled specimens were purified using 1.8X AMPure XP beads and eluted to 55 μl final volume (Beckman Coulter, Pasadena, CA, USA). Specimens that failed to amplify did not meet the criteria for library preparation and were excluded.

The pooled amplicons were diluted to 100-200 ng/ul for library preparation (Illumina DNA Prep kit) per the manufacturer’s instructions. Paired-end sequencing was performed on the Illumina NextSeq 550 platform using the v2.5, 300 cycle kit (Illumina, San Diego, CA).

**Supplementary Table 4.**
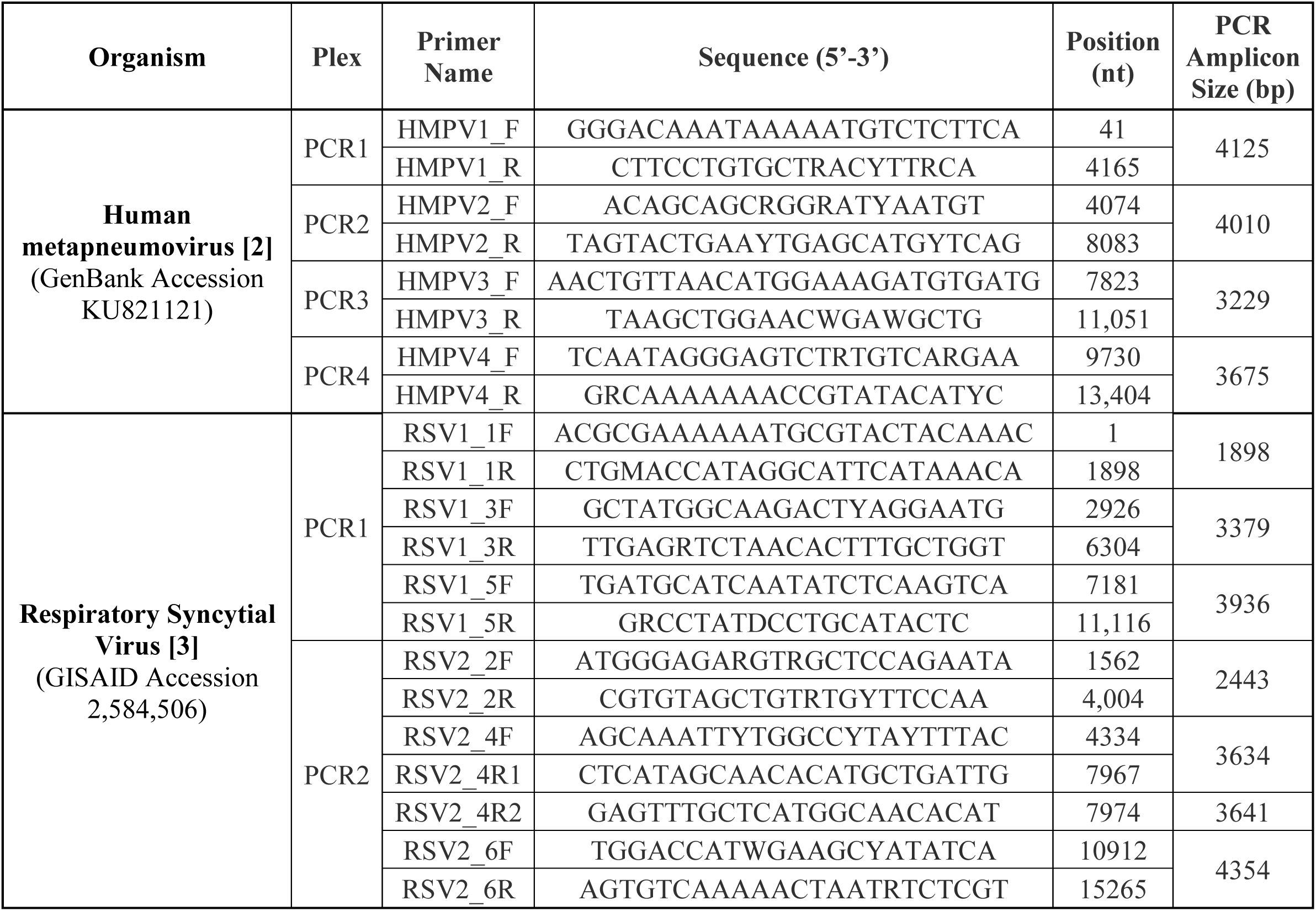
Information on primers used for multiplex PCR for human metapneumovirus (HMPV) and respiratory syncytial virus (RSV) (**adapted from Tullock et al 2021 and Dong et al 2023**)

